# Trajectory of weight regain after cessation of GLP-1 receptor agonists: a systematic review and nonlinear meta-regression

**DOI:** 10.1101/2025.06.09.25328726

**Authors:** Brajan Budini, Steven Luo, Martin Tam, Isabel Stead, Andrew Lee, Angelica Akrami, Antonio Vidal-Puig, Adrian Park

## Abstract

**Background:** Glucagon-like peptide 1 receptor agonists (GLP-1RAs) have emerged as breakthrough weight loss agents. However, discontinuation is common, and clinical trials have demonstrated significant weight regain following cessation. In this systematic review, we aimed to characterise the trajectory of weight regain after GLP-1RA cessation.

**Methods:** This systematic review and meta-regression analysis followed Cochrane and PRISMA guidelines. We searched MEDLINE, Embase, Cochrane Library, Scopus and Web of Science from inception to December 26, 2024 for randomised controlled trials and observational studies reporting weight outcomes after cessation of GLP-1RAs in adults with overweight or obesity. Weight regain was the primary outcome and was modelled using nonlinear regression. Secondary outcomes included HbA1c and systolic blood pressure. The study protocol is registered with PROSPERO (CRD420250631751).

**Findings:** We identified 44 relevant studies. Weight, HbA1c and systolic blood pressure consistently rebounded after cessation of GLP-1RAs. Six trials with 3,236 participants were included in the exponential recovery model. Weight regain was estimated to plateau at 75.6% (95% CI 68.5-82.7) of the weight lost on treatment. The rate constant was 0.0302 per week (95% CI 0.0204-0.0399), corresponding to a half-life of 23.0 weeks. At 1 year after cessation, an estimated 40.2% of the on-treatment weight loss remained. Most studies were assessed to have moderate risk of bias.

**Interpretation:** GLP-1RA cessation is associated with a predictable and decelerating pattern of weight regain, which appears to plateau below pre-treatment levels, suggesting that partial weight-loss benefit may persist long-term but is substantially attenuated.

**Funding:** None.

## Introduction

The increasing prevalence of obesity represents a major public health concern. Worldwide, more than one billion individuals are living with obesity^1^. At the individual level, obesity has been linked to poor health outcomes and is associated with comorbidities including type 2 diabetes, cardiovascular disease, and metabolic dysfunction-associated steatotic liver disease.

Weight loss of 5 to 10% can mitigate obesity-related health complications^2^. For patients with a body mass index (BMI) over 35 kg/m^2^, guidance^3^ recommends a greater weight loss of 15 to 20% for sustained improvement in comorbidity. Lifestyle modifications, primarily diet and exercise, have historically been the cornerstone of weight management. However, despite being widely recommended, lifestyle modification alone is generally ineffective^4^.

Until recently, pharmacologic options for weight loss were limited, with either poor efficacy or tolerability^5^. In the past few years, however, glucagon-like peptide 1 receptor agonists (GLP-1RAs) have emerged as highly effective weight loss drugs; clinical trials have demonstrated substantial weight losses of 15 to 20% with agents such as semaglutide and tirzepatide^6,7^. Patients treated with GLP-1RAs also see improvements in cardiometabolic parameters, including blood pressure, HbA1c, LDL and triglycerides^6^.

Despite the successes of GLP-1RAs, approximately half of patients who initiate them discontinue within the first year^8^, likely due to their considerable gastrointestinal side effects as well as limited access under insurance coverage policies and national prescribing guidelines.

Outcomes after discontinuing GLP-1RAs have been underexplored in the literature. One review^9^ reported weight regain after GLP-1RA cessation but did not examine the trajectory of weight over time. While clinical trials such as STEP 4^10^ and SURMOUNT-4^11^ have provided insights into post-cessation outcomes, additional relevant data exist across a wider body of trials with greater diversity in drugs and populations. Understanding the nature of weight regain could aid the development of strategies to maintain weight after cessation. Therefore, the aim of this review was to synthesise existing evidence and characterise the typical trajectory of weight regain following cessation of GLP-1RA treatment.

## Methods

### Search strategy and selection criteria

This systematic review and meta-regression followed the Cochrane Handbook for Systematic Reviews. Findings were reported according to PRISMA guidelines. The protocol is registered with PROSPERO (CRD420250631751). A systematic search was conducted by SL in MEDLINE, Embase, Cochrane Library, SCOPUS and Web of Science from their inception to Dec 26, 2024. We aimed to identify studies which tracked weight outcomes in patients following discontinuation of GLP-1RA therapy. A broad search strategy was used because post-cessation outcomes were often absent from abstracts despite being reported in full texts or supplements.

The search queries comprised variations of the term ‘glucagon-like peptide 1 receptor agonist’ in combination with keywords related to weight regain and treatment cessation. In addition, supplementary searches were conducted by SL in Google Search and Google Scholar to identify further relevant studies not included in the primary databases. Conference abstracts and other grey literature were included if they met inclusion criteria. Protocol papers and conference abstracts were followed up for full-text publications which, if eligible, were subsequently included in the review. Non-English studies and reviews were filtered out of search results. The complete search strategy is provided in the appendix (p 2).

Citations were imported into EndNote and duplicates were removed. Deduplicated citations were then uploaded to Rayyan for screening. Six authors conducted title and abstract screening and full-text screening. Each record was independently screened by two reviewers, and conflicts were resolved through discussion.

Studies were eligible for inclusion if they were randomised controlled trials (RCTs) or observational studies that reported weight outcomes following cessation of GLP-1RA treatment in adults with overweight or obesity, defined as a BMI of ≥25 kg/m^2^ or ≥23 kg/m^2^ for specific populations. Studies were required to have a treatment period of at least 8 weeks, a post-cessation follow-up period of at least 4 weeks and could not include co-treatment with other FDA-approved weight loss drugs. Case reports, protocol-only publications, reviews, editorials and non-English publications were excluded.

### Outcomes

The primary outcome was percentage weight regain, defined as the percentage of weight regained after treatment cessation relative to the weight lost at the end of treatment. This was calculated by using end-of-treatment weight loss data and the weight reported at any post-cessation time point available. We adopted this approach to normalise for differences in the absolute amount of weight lost, which varied between studies. Secondary outcomes included post-cessation changes in HbA1c and systolic blood pressure (SBP).

### Data analysis

Risk of bias was assessed using the Cochrane RoB 2 tool for RCTs and the Cochrane ROBINS-I tool for non-randomised studies. Risk of bias assessments were performed by BB, IS and SL, with each study being independently assessed by two reviewers. Conflicts were resolved through discussion.

Structured data extraction was performed using Google Sheets. Extracted variables included weight, HbA1c and SBP at baseline and follow-up at all available time points. Data values were independently extracted by two reviewers with discrepancies resolved by discussion. For each observation, we recorded the sample size, mean and standard error of the mean (SEM). Where data were presented graphically, we used WebPlotDigitizer to extract numerical values. If SEM was not reported directly, it was calculated from the confidence interval where available, or otherwise from the standard deviation and the sample size.

In this meta-regression, we aimed to characterise the trajectory of weight regain following GLP-1RA treatment cessation. To ensure reliable modelling, we filtered down the 44 included studies to include only those high-quality publications which comprehensively reported on post-cessation weight data. Our criteria required that studies be randomised, enrol a sufficient sample size in the treatment arm (n ≥ 100), report substantial on-treatment weight loss (≥ 3 kg) and have at least two post-cessation data points or one data point after 12 weeks. Two outlier studies were excluded to preserve comparability: one^12^ which focused on a population with type 2 diabetes, and another^13^ which investigated beinaglutide, a short-acting GLP-1RA with limited regulatory approval and markedly distinct pharmacokinetics^14^. Sensitivity analyses with these studies included showed little effect on the overall result. Ultimately, six studies^6,7,10–12,15^ were included in the meta-regression (table 1).

**Table 1:** Baseline characteristics for the trials included in the meta-regression analysis. Data are reported as mean (standard deviation). ^*^SURMOUNT-1 trialed multiple doses of tirzepatide; only data from the 15 mg group are presented here but all doses were included in the meta-regression. ^†^Treatment arm only. ^‡^All drugs were administered via subcutaneous injection. ^§^Percentage of weight lost at the end of the treatment period.

|  | STEP 1 | STEP 4 | STEP 10 | SURMOUNT-1* | SURMOUNT-4 | SCALE Obesity |
| --- | --- | --- | --- | --- | --- | --- |
| **Cohort size<sup>†</sup>** | 228 | 268 | 138 | 253 | 335 | 1505 |
| **GLP-1RA** | Semaglutide | Semaglutide | Semaglutide | Tirzepatide | Tirzepatide | Liraglutide |
| **Dosing<sup>‡</sup>** | 2.4 mg once weekly | 2.4 mg once weekly | 2.4 mg once weekly | 15 mg once weekly | 10 or 15 mg once weekly | 3 mg once daily |
| **Treatment length (weeks)** | 68 | 20 | 52 | 176 | 36 | 160 |
| **Follow-up length (weeks)** | 52 | 48 | 28 | 17 | 52 | 12 |
| **Age** | 48 (12) | 46 (12) | 53 (11) | 48 (12) | 48 (12) | 48 (12) |
| **Sex (% female)** | 66.7 | 76.5 | 72.5 | 63.6 | 70.7 | 75.8 |
| **BMI (kg/m<sup>2</sup>)** | 37.6 (7.0) | 38.4 (6.9) | 39.9 (6.6) | 39.2 (7.4) | 38.4 (6.6) | 38.8 (6.4) |
| **Weight (kg)** | 105.6 (21.8) | 107.2 (22.7) | 111.9 (21.5) | 108.6 (25.4) | 107.3 (22.3) | 107.5 (21.6) |
| **Change<sup>§</sup> (%)** | -17.1 | -11.0 | -13.9 | -22.9 | -20.0 | -6.1 |
| **HbA1c (%)** | 5.7 (0.3) | 5.7 (0.3) | 5.9 (0.3) | 5.8 (0.4) | 5.5 (0.4) | 5.8 (0.3) |
| **SBP (mmHg)** | 129 (14) | 127 (14) | 131 (15) | 125 (13) | 126 (13) | 125 (13) |

Due to significant variability in reported post-cessation time points between studies, we opted to model weight regain as a continuous trajectory using all available time points. In major trials assessing weight following cessation, namely STEP 1^6^, STEP 4^10^ and SURMOUNT-4^11^, we noted that weight regain resembled an exponential recovery curve approaching an asymptote. This observation is consistent with established first-order kinetic models of weight change, which predict rapid early changes followed by gradual plateauing as a new steady state is reached^16^. Therefore, we adopted an exponential recovery function for use in regression analysis.

Percentage weight regain was modelled as a function of time with the following formula:

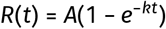

where *R*(*t*) is the percentage of weight regained at time *t* weeks, *A* is the asymptotic maximum weight regain, and *k* is the recovery rate constant. *A* was modelled with a fixed effect across all studies. *k* was modelled with fixed and study-level random effects to estimate the mean effect across all studies and account for between-study heterogeneity respectively. Model fit was assessed with root mean square error (RMSE) and Akaike information criterion (AIC). Alternative model specifications were tested but resulted in visually poorer fits with higher AIC. Inverse-variance weighting was applied using the SEM of each observation. Additionally, between-study heterogeneity in *k* was quantified as the variance of the random effects, *τ*^2^. Data analysis and visualisation were conducted using R (version 4.5.0) with the nlme and ggplot2 packages.

## Results

The study selection process is outlined in figure 1. The initial search yielded 5645 records. After deduplication and screening, 44 relevant studies were included in the review, comprising 33 RCTs and 11 observational studies.

**Figure 1:**
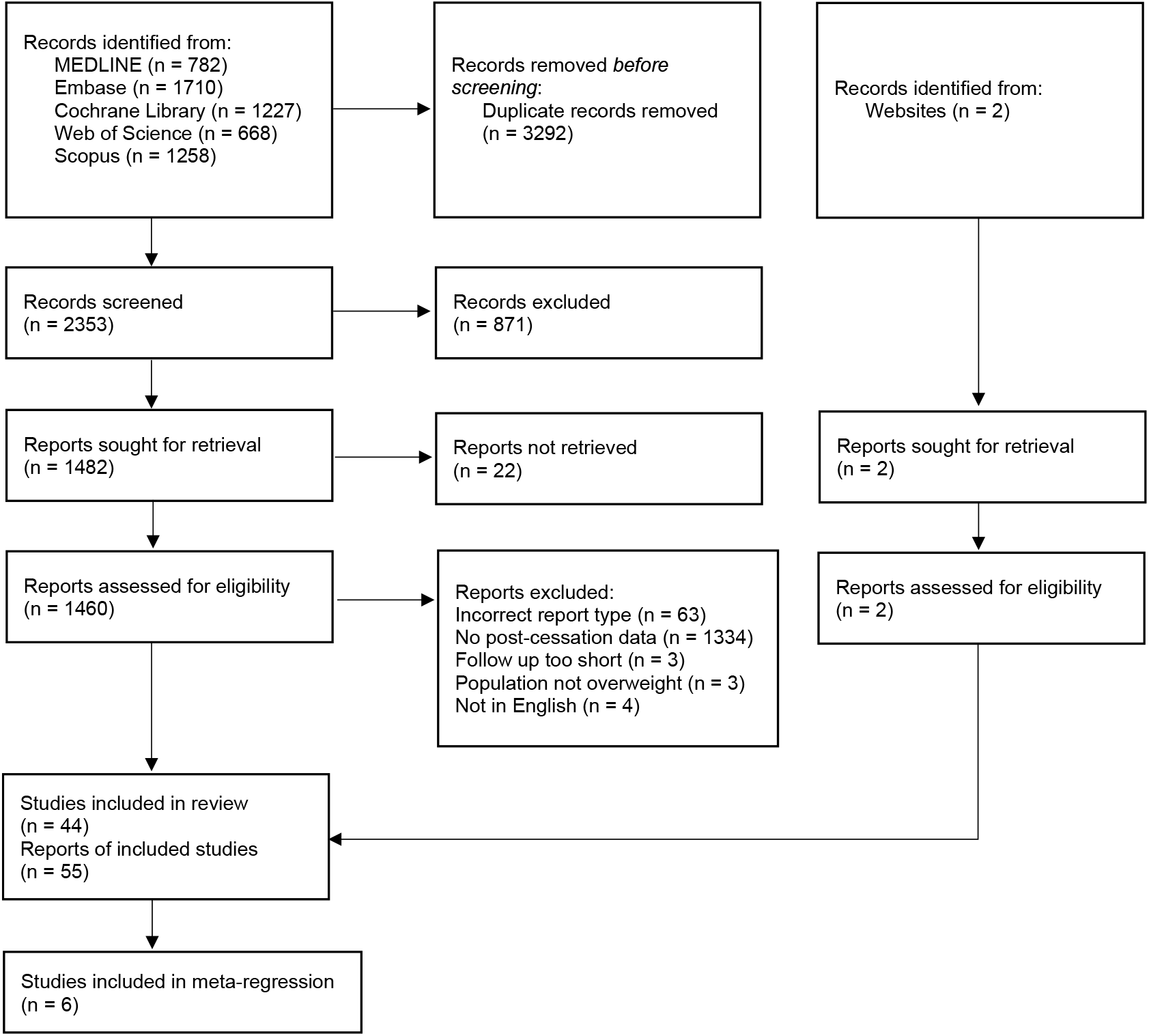
PRISMA flow diagram of included studies.

All studies reported post-cessation weight data in overweight and obese populations, with some focusing on comorbidities such as type 2 diabetes. Treatment durations ranged from 10 to 104 weeks and post-cessation follow-up durations spanned from 4 to 104 weeks. Interventions included liraglutide, semaglutide, tirzepatide, and other GLP-1RAs. A complete summary of the included studies is provided in the appendix (p 3).

Six RCTs were included in a mixed-effects exponential recovery regression to characterise the trajectory of weight regain. These trials included a total of 3,236 participants with longitudinal data on weight regain up to 52 weeks after GLP-1RA cessation. The baseline characteristics of these trials are summarised in table 1.

The estimated fixed effects were a maximum percentage weight regain (*A*) of 75.6% (95% CI 68.5-82.7) and a recovery rate constant (*k*) of 0.0302 per week (0.0204-0.0399), corresponding to a half-life of 23.0 weeks (95% CI 17.4–34.0). Model fit was acceptable with an RMSE of 7.60 percentage points based on fixed-effect predictions. Between-study heterogeneity of the recovery rate constant *k* was modest, with a *τ*^2^ of 9.77 × 10^−5^.

The fitted trajectories are shown in figure 2. The model estimated a rapid initial rate of weight regain which progressively slowed down and began to plateau at around 52 weeks, at which point the weight regain was 60% of the original weight loss or 79% of the estimated maximum weight regain. Weight regain trajectories appeared broadly similar across trials of liraglutide, semaglutide and tirzepatide.

**Figure 2:**
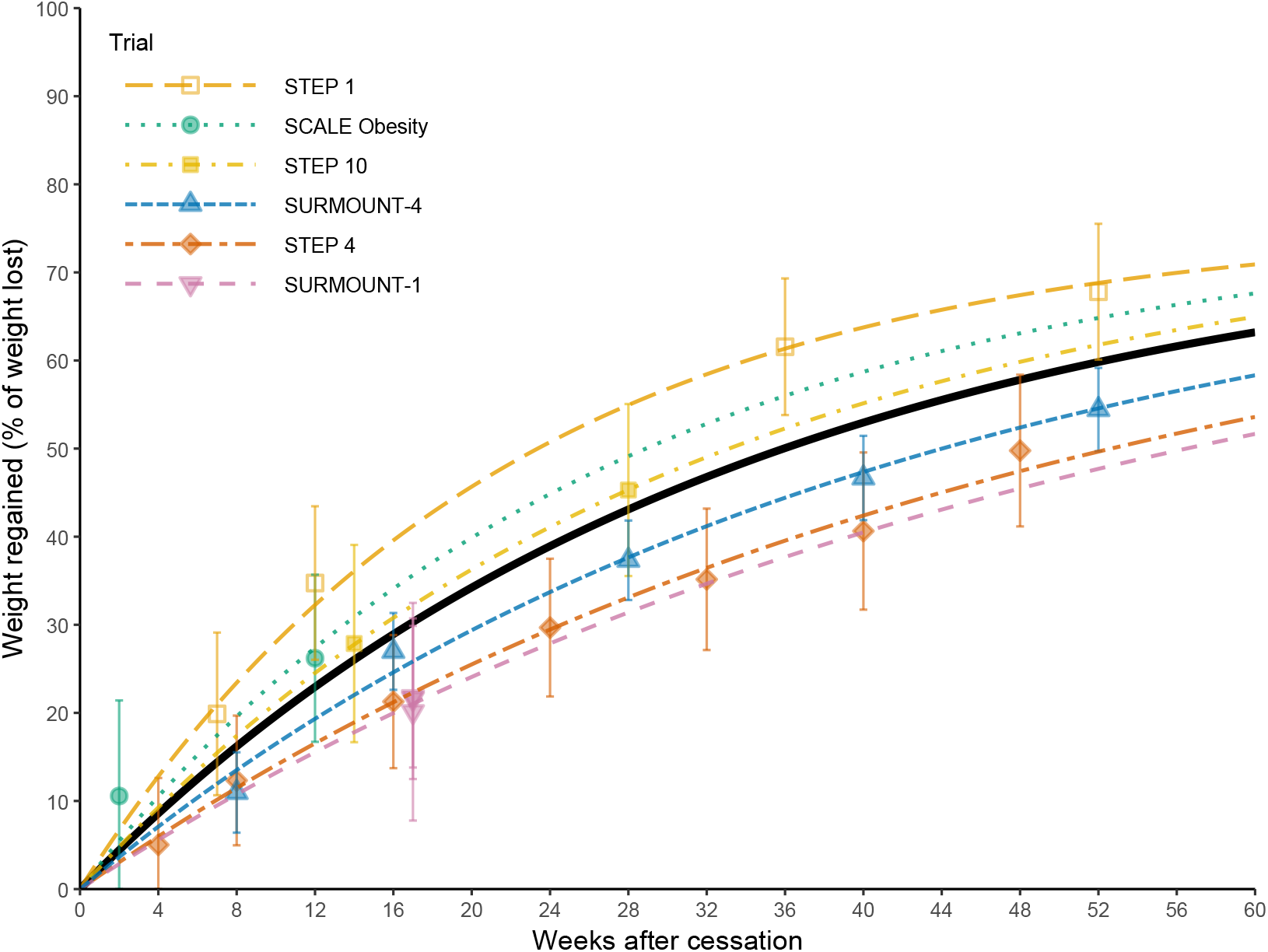
Fitted trajectories of weight regain after GLP-1RA cessation based on trial data. Regression was conducted with an exponential recovery function. Semaglutide was given in STEP 1^6^, STEP 4^10^ and STEP 10^15^; tirzepatide was given in SURMOUNT-1^7^ and SURMOUNT-4^11^; liraglutide was given in SCALE Obesity^12^. Black line represents the overall trajectory.

In our broader review, all studies reported weight reductions after treatment with GLP-1RAs, but the magnitude of weight loss ranged from around 1 to 20% of the pre-treatment baseline. This variation was largely driven by differences in drug efficacy—newer drugs including semaglutide and tirzepatide produced greater weight loss than older drugs such as exenatide and liraglutide.

Almost all studies observed some degree of weight regain after cessation of GLP-1RA treatment, although the extent varied significantly. Across studies with sufficient initial weight loss for meaningful interpretation, defined here as over 3 kg, the weight regained at or around 12 weeks post-cessation ranged from approximately 10 to 80% of the treatment-induced weight loss. Studies where participants lost little weight during treatment, defined here as less than 3 kg, were considered less reliable for interpretation. In one such study^17^, post-cessation weight exceeded the pre-treatment baseline weight, likely due to participants switching to insulin therapy, resulting in an apparent weight regain value of over 100%.

Post-cessation HbA1c was reported by 16 RCTs and five observational studies. HbA1c typically decreased by 0.5 to 1.5 percentage points while on GLP-1RA treatment, with some outliers falling outside this range. Virtually all studies which tracked this outcome observed post-cessation increases in HbA1c. In most cases, around half of the initial HbA1c reduction appeared to be regained by 8 to 12 weeks after treatment cessation. However, the extent and timing of this rebound varied significantly between studies. HbA1c rarely returned fully to pre-treatment baseline levels, even after one year off treatment.

Post-cessation SBP was reported by 15 RCTs and three observational studies. During treatment, SBP typically showed reductions ranging from around 1 to 10 mmHg, although some studies fell outside this range. After cessation of treatment, SBP typically rebounded towards the pretreatment baseline, with most studies reporting that around 70 to 80% of the initial SBP reduction was regained within 12 weeks.

Inadequate data and the lack of a clear or consistent temporal trajectory precluded further analysis of HbA1c and SBP.

Studies varied significantly in terms of the reporting of outcomes. Most trials were not explicitly designed to measure post-cessation outcomes and only extended data collection beyond treatment cessation for exploratory or safety reasons, typically for 4 to 12 weeks with a single data point taken at the end. Some trials, such as STEP 4^10^ and SURMOUNT-4^11^, were designed to measure post-cessation outcomes and often included longer follow-up durations of up to 52 weeks, with data collected at multiple intervening time points. This diversity in follow-up timing prompted us to choose a time-based regression approach rather than grouping data into discrete time windows.

Study populations showed heterogeneity in health conditions. 16 studies looked only at over-weight or obese patients with no specific comorbidities. 16 studies focused on patients with type 2 diabetes. Other comorbidities included metabolic dysfunction-associated steatotic liver disease, polycystic ovary syndrome, chronic kidney disease and chronic obstructive pulmonary disease. We were unable to determine whether comorbidities influenced the dynamics of post-cessation weight regain.

Some studies included interventions other than GLP-1RAs. Most studies mentioned some form of lifestyle counselling. Studies on patients with type 2 diabetes typically allowed for the use of standard diabetes medications such as metformin. Two studies^18,19^ prescribed an intensive low-calorie diet prior to liraglutide treatment, with liraglutide being used to maintain the already achieved weight loss. After cessation of liraglutide, weight appeared to return towards the preintervention baseline in a pattern broadly similar to that seen in other studies.

Three studies^20–22^ reported little to no weight regain after substantial on-treatment weight loss. Seier et al.^20^ implemented a dose tapering protocol and reported that patients’ mean weight did not increase after they reduced their dose of semaglutide. While the protocol allowed for complete cessation, many remained on some dose of semaglutide and it is unclear how many stopped entirely. In another study^21^, patients were maintained on a carbohydrate-restricted diet after cessation and experienced negligible weight regain. One study^22^ only reported categorical weight outcomes; while the distribution suggested no substantial changes in the average weight after drug cessation, details were limited, and the absence of mean values limited interpretation.

In one study of exenatide^23^, patients underwent two phases of treatment—52 weeks followed by 104 weeks—with a 12-week off-treatment period after each phase. Weight regain in the second off-treatment period was attenuated compared to the first, suggesting that GLP-1RA-induced weight loss may become more durable with prolonged exposure. However, we were unable to explore this hypothesis further due to insufficient data. Most studies were judged to have a moderate risk of bias, primarily due to the absence of pre-specified post-cessation outcomes in study protocols or analysis plans. Full risk of bias assessments are given in the appendix (p 6).

In summary, the majority of studies found partial weight regain after cessation of GLP-1RA therapy, with significant variation in magnitude and timing. Our regression model supported an exponential recovery trajectory, with a weight regain plateau at 76% of the original weight loss. Secondary outcomes of HbA1c and SBP generally showed rebound, but data were insufficient for modelling.

## Discussion

The findings of this review indicate that there is significant weight regain following cessation of GLP-1RAs. The meta-regression shows that after 1 year of treatment withdrawal, participants regain 60% of the weight they lost during treatment. The trajectory of regain is nonlinear and decelerating in nature, with an initial rapid recovery followed by a gradual tapering off and eventual plateauing. Notably, the estimated plateau is below the pre-treatment baseline weight, indicating that some beneficial effects may persist at the population level beyond 1 year after cessation, potentially indefinitely based on model extrapolation.

Weight regain poses a major challenge to the long-term efficacy of GLP-1RAs after treatment cessation, which is particularly relevant as around half of patients discontinue treatment within the first year^8^. Discontinuation is often due to gastrointestinal side effects, financial constraints, and limited access through insurance or prescribing guidelines. Despite this, public health bodies have yet to provide clear and coherent guidance on the use of GLP-1RAs in long-term weight management. In the UK, the National Institute for Health and Care Excellence (NICE) recommends that semaglutide should be prescribed for weight loss for a maximum of 2 years^24^, but no strict limit is set for tirzepatide^25^. This inconsistency highlights a blind spot in long-term obesity management as the benefits of GLP-1RAs risk being negated by subsequent weight regain.

While our model predicts considerable weight regain, it also suggests that 24% of the initial weight loss may be sustained long-term. This residual weight loss corresponds approximately to a 4 to 5% reduction in body weight relative to the pre-treatment baseline, assuming an initial weight loss of 15 to 20% as seen in trials of semaglutide and tirzepatide^6,7^. Notably, we observed that the residual weight loss was consistently greater than that seen in placebo groups. Clinically significant weight loss has been defined as a 5 to 10% reduction in body weight^2^, with the predicted residual weight loss of 4 to 5% falling just below this threshold. Nonetheless, GLP-1RAs may confer modest long-term metabolic benefits for some patients even after cessation.

If weight regain does indeed persist, this may be explained by sustained behavioural or physiological adaptations. By reducing appetite, GLP-1RAs may help facilitate the development of healthier eating habits, such as reduced portion sizes or improved nutritional quality, which may endure even after the pharmacological agent is removed. GLP-1RAs may also facilitate long-term physiologic adaptations such as altering hormone levels and hypothalamic resetting^26^.

When deprescribing GLP-1RAs, physicians and patients should be aware of the potential for weight regain and consider strategies to mitigate this risk. One possible strategy is individualised dose tapering. This approach was explored by Seier et al.^20^, who treated patients with semaglutide until they reached their target weight loss, then gradually reduced the dosage to the minimum dose that maintained their weight. This strategy may improve long-term tolerability, however patients may be unable to fully stop treatment. Another study^27^ is currently investigating whether weight regain after total discontinuation can be mitigated by interventions including tailored meals and a mobile application that tracks food intake and exercise.

Our results are subject to certain limitations and should be interpreted with caution. Most importantly, the trial data used to fit our model only extend to 52 weeks after cessation. No large-scale trials have reported weight regain data beyond this point; thus predictions made outside of this window are extrapolations. Longer-term trials are necessary to confirm that our predictions hold beyond one year. While we initially aimed to model trajectories of HbA1c and SBP following cessation, this proved infeasible due to inconsistent reporting and the lack of a clearly defined trajectory. As a result, insights into these parameters were limited, mainly confirming expected post-cessation rebound trends.

The topic of weight regain after GLP-1RA cessation has been largely underexplored in the literature. There are relatively few major trials investigating weight regain after cessation of GLP-1RAs; we only identified two large-scale RCTs that were designed to measure weight regain as a primary outcome: STEP 4^10^ and SURMOUNT-4^11^. Most of our included trials, including STEP 1^6^ and SURMOUNT-1^7^, measured weight regain as a secondary, often exploratory outcome; even these represent a small subset of all GLP-1RA trials, which generally do not report any post-cessation outcomes.

These results also raise questions about the composition of weight regained after cessation. Some studies suggest 40 to 60% of the weight lost on GLP-1RA treatment is lean mass^28^ and it is unclear whether the same proportion of lean mass is recovered during regain. If the regained weight has a higher proportion of fat, there could be negative impacts on cardiometabolic health. We noted a significant gap in the literature with regard to the composition of weight regain.

Our findings paint a nuanced picture regarding long-term post-GLP-1RA outcomes. Although a significant proportion of the weight is regained after cessation, the results of our modelling suggest that some benefit is maintained for at least 1 year, however there is likely significant variation at the individual level. As Seier et al.^20^ demonstrated, while some patients may require long-term therapy at the full dose for weight maintenance, others can taper their dose to a reduced maintenance level, and some may be able to discontinue therapy entirely whilst maintaining their desired weight. These considerations underscore the importance of a flexible, individualised approach to weight management when using GLP-1RAs. Blanket guidelines such as NICE’s 2 year limit for semaglutide may fail to accommodate patients’ individual needs and risk undermining long-term effectiveness.

The landscape of weight loss medications is rapidly evolving as new agents are brought to market. Although included here as a GLP-1RA, tirzepatide is a dual agonist that also targets the glucose-dependent insulinotropic polypeptide (GIP) receptor, likely explaining its superior efficacy^29^. Even more potent agents are on the horizon. Currently undergoing clinical trials is retatrutide, a triple agonist of the GLP-1, GIP and glucagon receptors, which has produced weight loss of 24%^30^. If the weight dynamics in this review hold for newer agents, greater on-treatment efficacy may translate to sizeable residual weight loss even after discontinuation.

In the face of high discontinuation rates and policy-imposed treatment duration limits, our findings underscore the need for an urgent re-evaluation of prescribing guidelines. Individualised cessation strategies and long-term support are essential to sustain treatment benefits and prevent the reversal of clinical progress. As newer and more potent agents emerge, understanding the trajectory of discontinuation outcomes will be pivotal in shaping future obesity care.

## Data Availability

Data for this study were extracted from the published literature. The dataset is available on request.

## Contributors

BB and SL conceived the study and co-wrote the first draft of the manuscript. BB, SL, MT and AA co-designed the study. SL conducted the systematic search. AA, AL, BB, IS, MT and SL conducted literature screening. AL, BB, IS and SL carried out data extraction. SL performed the data analysis. BB, IS and SL conducted risk of bias assessments. AVP and AP supervised the project, aided data interpretation and contributed to the final manuscript. All authors had the final responsibility for the decision to submit for publication.

## Acknowledgments

ChatGPT was used to verify grammar, refine the English phrasing in our text and for coding assistance in data analysis. All output was manually reviewed and the authors take full responsibility for the accuracy and quality of the publication.

