## Supplementary material for "Trajectory of weight regain after cessation of GLP-1 receptor agonists: a systematic review and nonlinear meta-regression"

**Table of contents**


Table S1. Search terms 2
Table S2. Summary of included studies 3
Figure S1. Risk of bias assessments 6
References 9

**Table S1. Search terms**

Ovid MEDLINE(R) and Epub Ahead of Print, In-Process,
In-Data-Review & Other Non-Indexed Citations, Daily and Versions
<1946 to December 26, 2024>

1 exp Glucagon-Like Peptide 1/ or (glucagon-like peptide 1 or GLP-1 or GLP1).ti,ab,kw,kf.

2 (agonist* or analog*).ti,ab,kw,kf.

3 (GLP-1RA* or GLP1RA*).ti,ab,kw,kf.

4 (liraglutide or semaglutide or tirzepatide or exenatide or lixisenatide or albiglutide or dulaglutide or efpeglenatide or beinaglutide or taspoglutide or pegapamodutide or mazdutide or retatrutide or danuglipron or orforglipron).ti,ab,kw,kf.

5 (1 and 2) or 3 or 4

6 (weight adj3 (gain* or regain* or los* or maint* or chang* or difference or increas* or decreas* or reduc* or declin* or trajectory or control* or normali* or fluctuat* or r?se* or rebound* or recover* or restor* or relaps*)).ti,ab,kw,kf.

7 randomized controlled trial.pt. or (random* and placebo).ti,ab,kw,kf.

8 week*.ti,ab,kw,kf.

9 7 and 8

10 (cohort or observational or retrospective or prospective).ti,ab,kw,kf.

11 (withdraw* or discontin* or ceas* or cessation or deprescri* or stop* or termin* or finish* or paus* or "not continu*" or suspend* or conclud*).ti,ab,kw,kf.

12 10 and 11

13 5 and 6 and (9 or 12)

**Table S2. Summary of included studies**

| Study | Design | Comorbidities (except obesity) | Total participants (all arms) | GLP-1RA | Cotreatments | Treatment length (weeks) | Post-treatment follow up length (weeks) |
|---|---|---|---|---|---|---|---|
| Altintas Dogan et al. (2022)[1] | RCT | COPD | 40 | liraglutide | - | 40 | 4 |
| Apperloo et al. (2025)[2] | RCT | CKD | 101 | semaglutide | ACE inhibitors, SGLT2 inhibitors | 24 | 4 |
| Armstrong et al. (2016)[3] | RCT | MASH | 52 | liraglutide | lifestyle advice | 48 | 12 |
| Aronne et al. (2024)[4] | RCT | - | 670 | tirzepatide | lifestyle advice | 36 | 52 |
| Asano et al. (2023)[5] | RCT | T2DM | 16 | cotadutide | - | 10 | 4 |
| Barnett et al. (2007)[6] | RCT (crossover) | T2DM | 138 | exenatide | metformin/sulfonylurea, insulin glargine after stopping exenatide | 16 | 16 |
| Bartelt et al. (2024)[7] | retrospective cohort | - | 38,007 | semaglutide, liraglutide | - | variable | 52 |
| Bunck et al. (2011)[8] | RCT | T2DM | 69 | exenatide | metformin | 52, 104 | 12,12 |
| Chen et al. (2024)[9] | RCT | - | 427 | beinaglutide | lifestyle advice | 16 | 12 |
| D'Alessio et al. (2014)[10] | RCT (crossover) | T2DM | 978 | liraglutide | metformin ± sulfonylureas, insulin glargine after stopping liraglutide | 24 | 24 |
| Davies et al. (2015)[11] | RCT | T2DM | 846 | liraglutide | lifestyle advice + metformin ± sulfonylureas, glitazone | 56 | 12 |
| Dusilová et al. (2024)[12] | RCT (crossover) | MASLD | 16 | semaglutide | dietary intervention continued after stopping semaglutide | 16 | 16 |
| Enebo et al. (2021)[13] | RCT | - | 96 | semaglutide | cagrilinitide or placebo | 20 | 5 |

| | | | | | | | |
|---|---|---|---|---|---|---|---|
| Ferjan et al. (2017)[14] | RCT | PCOS | 24 | liraglutide | metformin or metformin + sitagliptin after stopping liraglutide | 12 | 12 |
| Ferrari et al. (2020)[15] | retrospective cohort | - | 93 | liraglutide | lifestyle advice | variable | variable |
| Fineman et al. (2011)[16] | RCT | T2DM | 107 | exenatide | metformin + lifestyle advice | 15 | 12 |
| Frias et al. (2022)[17] | RCT | T2DM | 406 | efpeglenatide | lifestyle advice | 56 | 6 |
| Garcia de Lucas and Olalla Sierra (2017)[18] | prospective cohort | T2DM | 13 | liraglutide, lixisenatide, exenatide | canagliflozin | 36 | 26 |
| Gibbons et al. (2021)[19] | RCT (crossover) | T2DM | 15 | semaglutide | metformin + lifestyle advice | 12, 12 | 8, 8 |
| Jastreboff et al. (2025)[20] | RCT | - | 1,032 | tirzepatide | lifestyle advice | 176 | 17 |
| Jensen et al. (2024)[21] | RCT | - | 195 | liraglutide | low-calorie diet prior to starting liraglutide + lifestyle advice during treatment | 52 | 52 |
| Jensterle et al. (2024)[22] | prospective cohort | PCOS | 25 | semaglutide | metformin + lifestyle advice | 16 | 104 |
| Ji et al. (2023)[23] | RCT | - | 248 | mazdutide | - | 24 | 12 |
| Khoo et al. (2019)[24] | RCT | MASLD/MASH | 30 | liraglutide | lifestyle advice | 26 | 26 |
| Kubota et al. (2023)[25] | prospective cohort | T2DM | 9 | tirzepatide | - | 52 | 104 |
| Lau et al. (2021)[26] | RCT | - | 706 | liraglutide | lifestyle advice | 26 | 6 |
| le Roux et al. (2017)[27] | RCT | prediabetes | 2,254 | liraglutide | lifestyle advice | 160 | 12 |
| McGowan et al. (2024)[28] | RCT | prediabetes | 207 | semaglutide | lifestyle advice | 52 | 28 |
| McInnes et al. (2023)[29] | RCT | T2DM | 160 | lixisenatide | insulin glargine + metformin + lifestyle advice | 12 | 52 |

| | | | | | | | |
|---|---|---|---|---|---|---|---|
| McKenzie and Athinarayanan (2024)[30] | retrospective cohort | T2DM | 308 | unspecified | carbohydrate restricted nutrition therapy continued after stopping GLP-1RA | variable | 52 |
| Montvida et al. (2017)[31] | retrospective cohort | T2DM | 66,583 | unspecified | - | variable | variable |
| O'Neil et al. (2018)[32] | RCT | - | 957 | semaglutide, liraglutide | lifestyle advice | 52 | 7 |
| Punthakee et al. (2024)[33] | RCT | T2DM | 159 | liraglutide | insulin degludec + metformin + lifestyle advice | 16 | 52 |
| Rosenstock et al. (2010)[34] | RCT | - | 152 | exenatide | lifestyle advice | 24 | 4 |
| Rubino et al. (2021)[35] | RCT | - | 803 | semaglutide | lifestyle advice | 20 | 48 |
| Sanyal et al. (2024)[36] | RCT | MASLD | 98 | retatrutide | lifestyle advice | 48 | 4 |
| Seier et al. (2025)[37] | prospective cohort | - | 2,694 | semaglutide | weight-management programme | variable | 26 |
| Siskind et al. (2020)[38] | RCT | clozapine-associated obesity | 28 | exenatide | clozapine | 24 | 52 |
| Svensson et al. (2019)[39] | RCT | clozapine-associated obesity | 103 | liraglutide | clozapine/olanzapine | 16 | 52 |
| Varanasi et al. (2011)[40] | retrospective cohort | T2DM | 141 | exenatide | lifestyle advice | variable | 26 |
| Wadden at al. (2013)[41] | RCT | - | 422 | liraglutide | low-calorie diet prior to starting liraglutide | 56 | 12 |
| Wilding et al. (2022)[42] | RCT | - | 327 | semaglutide | lifestyle advice | 68 | 52 |
| Yu et al. (2022)[43] | retrospective cohort | - | 157 | liraglutide | - | variable | variable |
| Zhou et al. (2023)[44] | prospective cohort | T2DM | 98 | liraglutide | lifestyle advice | 12 | variable |

#### Figure S1. Risk of bias assessments

A. Randomised controlled trials, weight

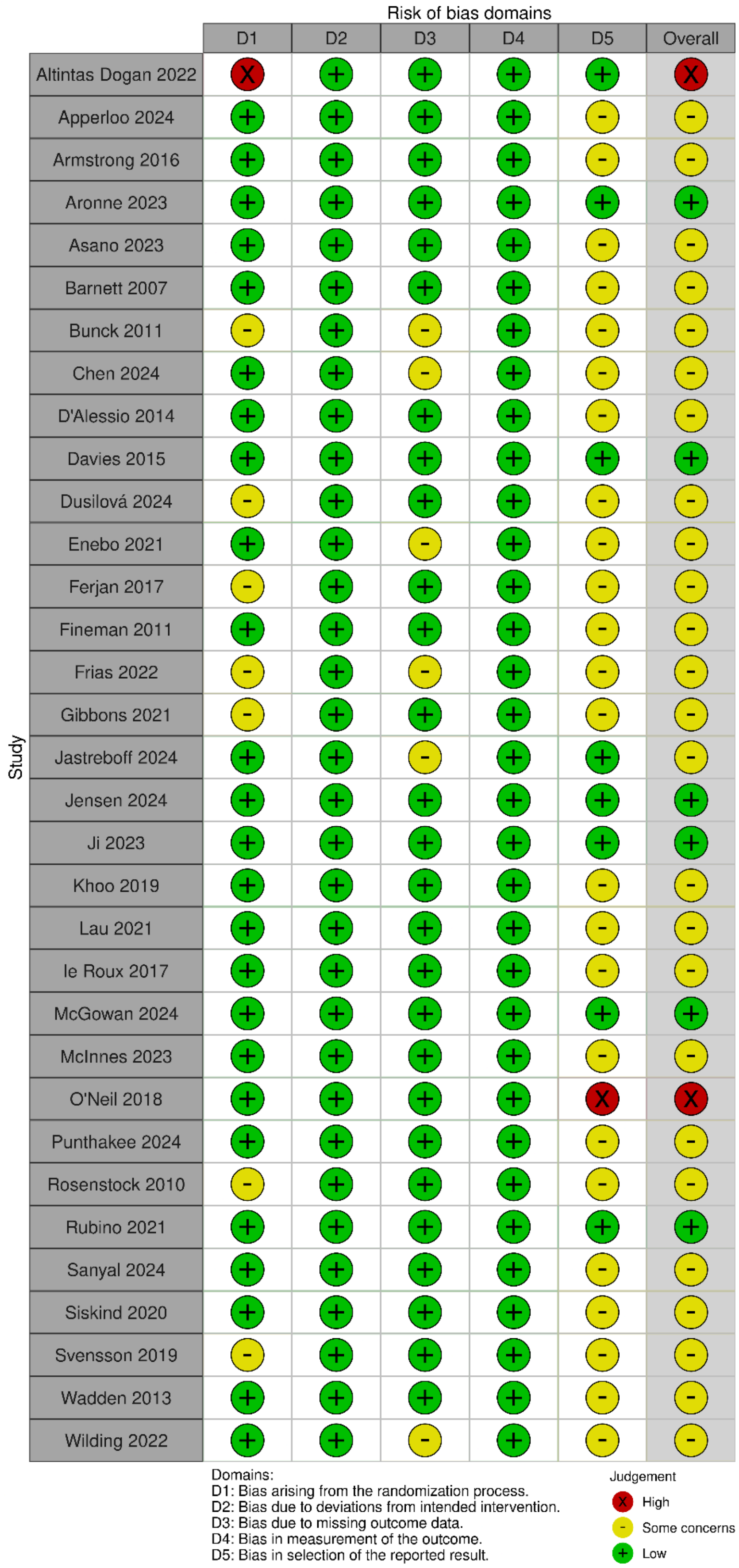

#### B. Randomised controlled trials, HbA1c

Risk of bias domains

| Study | D1 | D2 | D3 | D4 | D5 | Overall |
|---|---|---|---|---|---|---|
| Apperloo 2024 | + | + | - | + | - | - |
| Aronne 2023 | + | + | + | + | + | + |
| Asano 2023 | + | + | + | + | - | - |
| Barnett 2007 | + | + | + | + | - | - |
| Bunck 2011 | - | + | - | + | - | - |
| D'Alessio 2014 | + | + | + | + | - | - |
| Fineman 2011 | + | + | + | + | - | - |
| Frias 2022 | - | + | - | + | - | - |
| Jastreboff 2024 | + | + | - | + | + | - |
| Jensen 2024 | + | + | + | + | + | + |
| Armstrong 2016 | + | + | + | + | - | - |
| McGowan 2024 | + | + | + | + | + | + |
| Punthakee 2024 | + | + | + | + | - | - |
| Siskind 2020 | + | + | X | + | - | X |
| Svensson 2019 | - | + | + | + | - | - |
| Wilding 2022 | + | + | - | + | - | - |

Domains:
D1: Bias arising from the randomization process.
D2: Bias due to deviations from intended intervention.
D3: Bias due to missing outcome data.
D4: Bias in measurement of the outcome.
D5: Bias in selection of the reported result.

Judgement
X High
- Some concerns
+ Low

#### C. Randomised controlled trials, systolic blood pressure

Risk of bias domains

| Study | D1 | D2 | D3 | D4 | D5 | Overall |
|---|---|---|---|---|---|---|
| Apperloo 2024 | + | + | + | + | - | - |
| Armstrong 2016 | + | + | + | + | - | - |
| Aronne 2023 | + | + | + | + | + | + |
| Davies 2015 | + | + | + | + | - | - |
| Jastreboff 2024 | + | + | - | + | + | - |
| Jensen 2024 | + | + | + | + | + | + |
| Ji 2023 | + | + | + | + | + | + |
| le Roux 2017 | + | + | + | + | - | - |
| McGowan 2024 | + | + | + | + | + | + |
| Rubino 2021 | + | + | + | + | + | + |
| Siskind 2020 | + | + | + | + | - | - |
| Svensson 2019 | - | + | + | + | - | - |
| Wilding 2022 | + | + | - | + | - | - |

Domains:
D1: Bias arising from the randomization process.
D2: Bias due to deviations from intended intervention.
D3: Bias due to missing outcome data.
D4: Bias in measurement of the outcome.
D5: Bias in selection of the reported result.

Judgement
- Some concerns
+ Low

### D. Non-randomised studies, weight

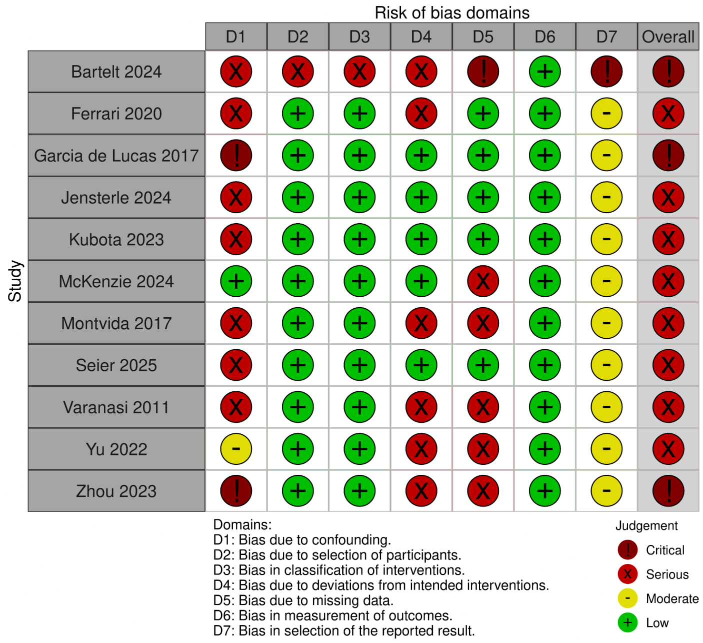


### E. Non-randomised studies, HbA1c

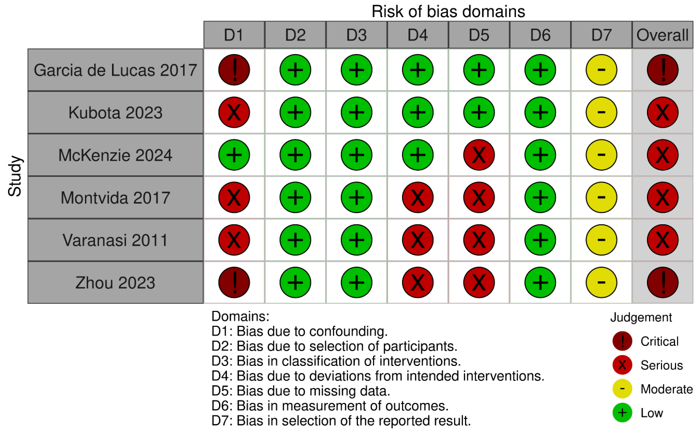


### F. Non-randomised studies, systolic blood pressure

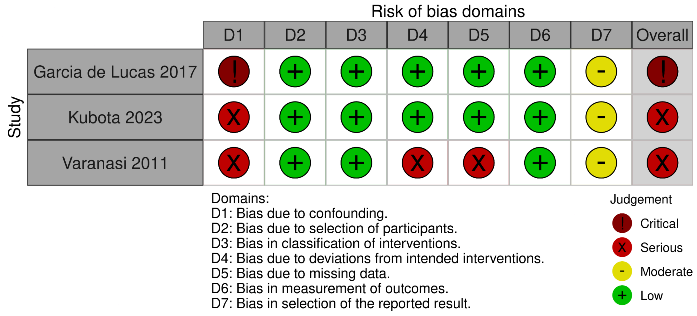

23. Ji L, Jiang H, Cheng Z, Qiu W, Liao L, Zhang Y, et al. A phase 2 randomised controlled

trial of mazdutide in Chinese overweight adults or adults with obesity. Nat Commun. 2023 Dec 14;14(1):8289.
